# PvDBPII-Matrix M elicits polyfunctional antibodies that limit parasite growth in a challenge trial

**DOI:** 10.1101/2023.08.01.23293515

**Authors:** Francisco J. Martinez, Michael White, Micheline Guillotte-Blisnick, Christèle Huon, Alix Boucharlat, Fabrice Agou, Patrick England, Jean Popovici, Mimi M. Hou, Sarah E. Silk, Jordan R. Barrett, Carolyn M. Nielsen, Jenny M. Reimer, Paushali Mukherjee, Virander S. Chauhan, Angela M. Minassian, Simon J. Draper, Chetan E. Chitnis

## Abstract

The receptor-binding domain, region II, of *Plasmodium vivax* Duffy binding protein (PvDBPII) binds the Duffy antigen on reticulocytes to mediate invasion. A heterologous vaccine challenge trial recently showed that a delayed dosing regimen with recombinant PvDBPII SalI formulated with adjuvant Matrix-M^TM^ reduced the *in vivo* parasite multiplication rate (PMR) challenged with the *P. vivax* Thai isolate PvW1. We describe extensive analysis of the polyfunctional antibody responses elicited by PvDBPII immunization and identify immune correlates for PMR reduction. A classification algorithm identified antibody features that contribute significantly to PMR reduction. These included antibody titre, receptor-binding inhibitory titre, dissociation constant for PvDBPII-antibody interaction, complement C1q and Fc gamma receptor binding and specific IgG subclasses. These data suggest that multiple immune mechanisms elicited by PvDBPII immunization are associated with protection. The identified immune correlates could guide the development of an effective vaccine for *P. vivax* malaria. Importantly, all the polyfunctional antibody features that correlated with protection cross-reacted with both PvDBPII SalI and PvW1 variants, suggesting that immunization with PvDBPII should protect against diverse *P. vivax* isolates.

## Introduction

*Plasmodium vivax* accounts for majority of malaria cases outside sub-Saharan Africa, where *P. falciparum* is more predominant (1). For many decades, *P. vivax* malaria was considered to be a ‘benign’ infection. However, recent studies have reported significant incidence of severe symptoms in *P. vivax* malaria (2,3). Ineffective control of *P. vivax* could pose an important public health threat to communities in which *P. vivax* is endemic or at risk of emerging. Current efforts to control malaria are less effective in reducing *P. vivax* compared to *P. falciparum* due to the unique biology of *P. vivax* (4). For example, the dormant liver stage of *P. vivax* known as the hypnozoite can reactivate and cause blood stage infection weeks, months or even years after the initial infection. This dormant stage, which cannot be detected, greatly contributes to *P. vivax* prevalence (5). In addition, *P. vivax* gametocytes appear early in the blood stage, so the parasite can be transmitted even before the first symptoms appear and treatment is administered to the patient (6). An effective vaccine that can prevent *P. vivax* infection, reduce blood stage replication and protect against disease could greatly help efforts to control and eventually eliminate *P. vivax*.

During the blood stage, the invasive form of *P. vivax* known as the merozoite repeatedly infects and multiplies within reticulocytes causing the clinical symptoms of malaria. The invasion of reticulocytes is mediated by the interaction of the *P. vivax* Duffy binding protein (PvDBP) and the Duffy antigen receptor for chemokines (DARC) (7,8). The amino-terminal cysteine-rich region II of PvDBP (PvDBPII), serves as the binding domain of this invasion ligand (9). Importantly, Duffy negative individuals remain largely resistant to *P. vivax* infection (7). Numerous reports indicate PvDBPII is highly polymorphic (10–12), suggesting that it is under intense immune pressure. However, the binding residues in PvDBPII that make contact with DARC are highly conserved (13–15). The binding residues, which include positively charged as well as hydrophobic amino acids, assemble on the PvDBPII surface to form a receptor recognition site that is fully exposed to neutralization by antibodies (15). Moreover, since the binding residues are conserved, antibodies that target them are predicted to be strain-transcending and capable of neutralizing diverse *P. vivax* strains. Indeed, upon natural exposure a small percentage of individuals develop high titre binding inhibitory antibodies against PvDBPII that are cross-reactive (16,17). Importantly, the presence of such high titre binding inhibitory antibodies against PvDBPII is associated with reduced risk of *P. vivax* infection (16,17). However, the low frequency of individuals in the population with such binding inhibitory antibodies indicates that natural exposure is not effective at eliciting protective anti-PvDBPII antibodies.

Pre-clinical studies demonstrated that immunization of animals with recombinant PvDBPII readily elicits high titre binding inhibitory antibodies that cross-react with multiple variants (18,19). A vaccine based on PvDBPII could thus potentially elicit binding inhibitory antibodies that limit blood-stage growth of diverse *P. vivax* isolates. To date, two delivery platforms developed for PvDBPII have been tested in clinical trials. These include recombinant PvDBPII protein formulated with adjuvants such as glucopyranosyl lipid adjuvant-stable emulsion (GLA-SE) (20) and PvDBPII delivered by two viral vectors, namely, replication deficient chimpanzee adenovirus serotype 63 (ChAd63) for priming followed by modified vaccinia virus Ankara (MVA) for boosting (21). The vaccine candidate PvDBPII/GLA-SE was shown to be safe and no related adverse events were associated with the vaccine (20). In addition, PvDBPII-specific antibodies were elicited and shown to inhibit DARC-binding by diverse PvDBPII variants (20). In a Phase I trial with UK adult volunteers, the viral-vectored PvDBPII showed similar safety and immunogenicity results (21). Recently, PvDBPII protein reformulated in Matrix-M^TM^ adjuvant (PvDBPII/Matrix-M^TM^) from Novavax, and PvDBPII expressed by the same viral vectors, ChAd63/MVA, were tested in parallel in a Phase I/IIa blood stage challenge clinical trial using a heterologous *P. vivax* isolate to evaluate efficacy (22). Interestingly, volunteers who were vaccinated with PvDBPII/Matrix-M^TM^ in a delayed dosing schedule showed significant reduction in the parasite multiplication rate (PMR) compared to unvaccinated volunteers. None of the other groups in the trial, including PvDBPII delivered by viral vectors, showed any reduction in PMR. This result provided proof-of-concept that immunization with PvDBPII can elicit immune responses that can impair *in vivo* growth of a heterologous *P. vivax* strain. Here, we report extended analyses of the antibody responses including functional analysis of anti-PvDBPII antibodies, PvDBPII-antibody binding kinetics, recognition of native PvDBP in *P. vivax* schizonts as well as binding to C1q and Fc gamma receptors, to identify immune correlates of protection following immunization with vaccine candidates based on PvDBPII.

## Results

### Functional analysis of antibody response to PvDBPII in a challenge trial

A schematic describing the challenge trials to evaluate efficacy of PvDBPII administered using two approaches for vaccine antigen delivery, namely, recombinant PvDBPII SalI protein adjuvanted with Matrix-M^TM^, PvDBPII/Matrix-M^TM^, and PvDBPII SalI delivered by viral-vectors, ChAd63 followed by MVA (ChAd63/PvDBPII and MVA/PvDBPII) (22) is shown in Supplementary Figure S1.

As described earlier (22), volunteers received priming immunizations with PvDBPII/Matrix-M^TM^ and ChAd63/PvDBPII in January 2020. The trial was halted in March 2020 due to the Covid-19 pandemic. Twelve volunteers in the PvDBPII/Matrix-M^TM^ arm (n = 12) received the priming dose and one booster dose of PvDBPII/Matrix-M^TM^ at 1 month whereas ten volunteers from the viral vectors arm (n = 10) had only received the priming dose of ChAd63/PvDBPII by the time the trial was put on halt in March 2020. The trial was restarted in April 2021 by which time several volunteers had dropped out of the trial. Six volunteers who received the first 2 monthly doses of the recombinant PvDBPII/Matrix-M^TM^ in January and February 2020 (days 0 and 28), got the final boost after 14 months (day 440) and underwent Controlled Human Malaria Infection (CHMI) with *P. vivax* blood-stage parasites (clonal strain PvW1) (23) in May 2021. We refer to this group as 79A. Two volunteers, who received the ChAd63/PvDBPII prime in January 2020 (day 0), received an additional ChAd63/PvDBPII followed by MVA/PvDBPII at 17 and 19 months (days 530 and 586), respectively. We designated this group as 71B’. New volunteers were enrolled in the trial and four of them received PvDBPII/Matrix-M^TM^ in a regimen of 0, 1 and 2 months (days 530, 558 and 586) and three others received ChAd63/PvDBPII followed by MVA/PvDBPII scheduled at 0 and 2 months (days 530 and 586). These short regimen groups (with vaccines given as originally intended) are referred to as groups 79B and 71B, respectively. Volunteers from 79B, 71B’ and 71B groups underwent CHMI in October 2021. Regardless of the vaccine regimen, all volunteers underwent CHMI 2-4 weeks after the last immunization. Seven individuals who did not received any vaccine were included as the control group in the CHMI conducted in May 2021 (69C group) and four non-vaccinated volunteers underwent CHMI in October 2021 (69D group). Both CHMI challenges involved infection with blood stage parasites of the heterologous *P. vivax* monoallelic isolate W1 (or PvW1) (23). Parasite growth was evaluated in immunized and control unvaccinated groups by RT-PCR at different time points during the CHMI to determine vaccine efficacy (22).

As reported earlier (22), only the 79A group showed a significant reduction of the PMR compared to control groups (median PMR of 3.2 for 79A group and median PMR of 6.8 for controls, p = 0.002). This represented a 53% reduction of the median PMR in 79A group compared to controls. No other vaccine group had a significant impact on the PMR. The PMR data from the challenge trial (22) is summarized for clarity in Supplementary Table S1.

ELISA antibody titres for recognition of the homologous PvDBPII SalI domain by sera from immunized volunteers were determined through the course of the trial (Figure 1A). Anti-PvDBPII antibody titres peaked 2 weeks after final boost in all vaccinated volunteers. The antibody titres of sera in the 79A group (PvDBPII/Matrix-M^TM^) increased significantly after the delayed second boost at day 440. In case of group 79B (PvDBPII/Matrix-M^TM^), while there was a significant increase in antibody titres after the first monthly boost, there was no significant increase after the second monthly boost. The anti-PvDBPII antibody titres in group 79A, which was the only group to show a significant PMR reduction, remained stable after second boost and challenge with only a marginal decline over a 4-month period after the delayed second boost (Figure 1A). In the groups immunized with viral-vectored PvDBPII, the anti-PvDBPII antibodies reached a peak after MVA boost prior to challenge (Figure 1A).

**Figure 1.**
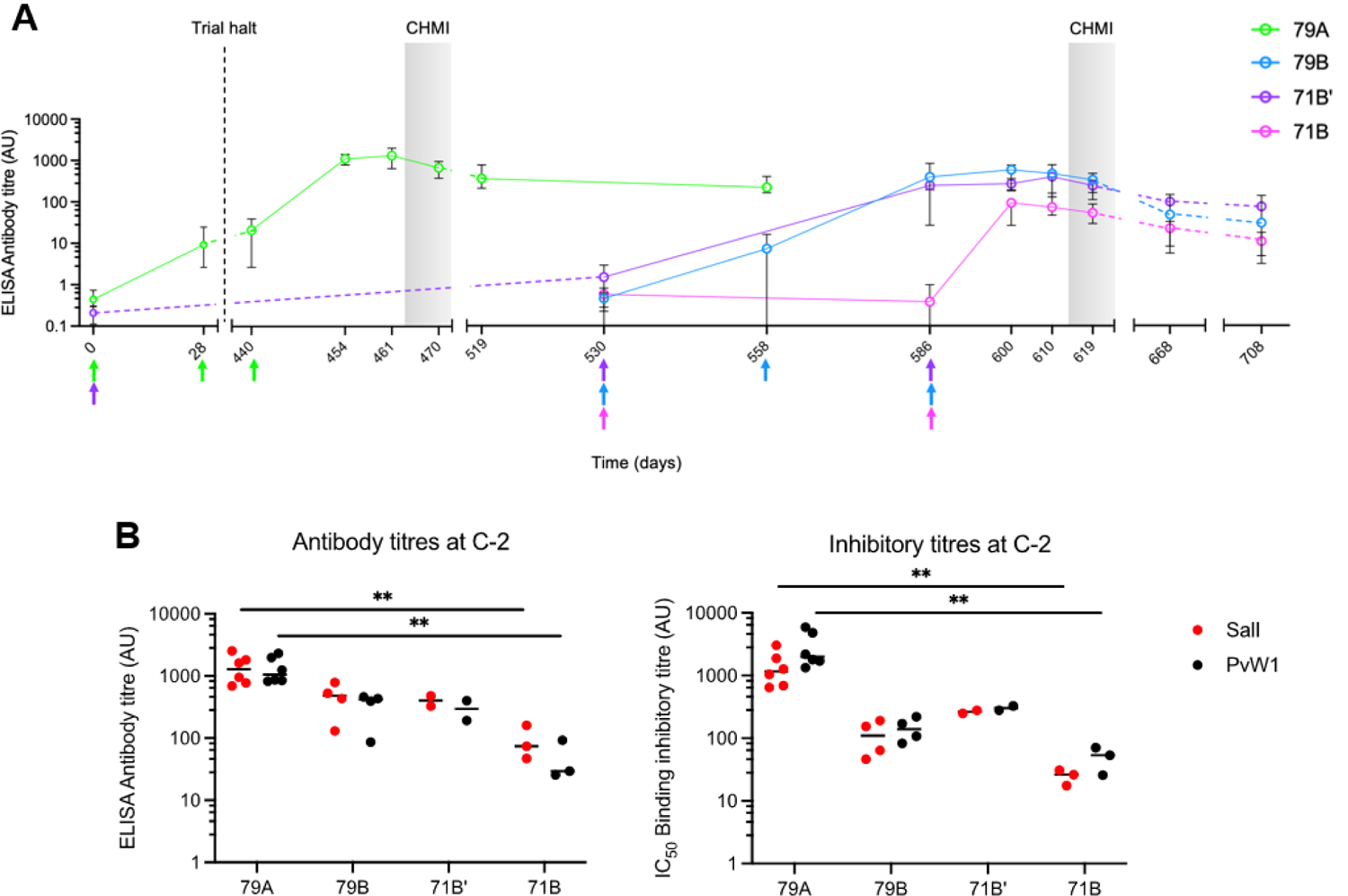
Antibody and binding inhibitory responses after immunization with protein-in-adjuvant or viral-vectored PvDBPII vaccines. (**A**) ELISA antibody titres for recognition of PvDBPII SalI over the course of the trial reported as median and range for each vaccine group are shown. Vertical dashed line indicates the trial halt. Colored arrows show the time points of the immunizations. Shaded grey areas represent the two CHMIs. (**B**) ELISA antibody titres and binding inhibitory titres specific to PvDBPII variants SalI and PvW1 for all volunteers at C-2. Medians are shown in horizontal bars, **p < 0.01, Kruskal–Wallis test with Dunn’s correction for multiple comparisons.

Next, we compared the antibody responses at C-2, 2 days before challenge, against PvDBPII PvW1, the heterologous PvDBPII domain from the *P. vivax* strain used for CHMI and the homologous PvDBPII SalI domain used for immunization. Sera from the immunized volunteers had similar ELISA recognition titres for PvDBPII SalI and PvW1 in all the groups (Figure 1B). Also, at C-2, the antibody titres of group 79A for recognition of PvDBPII were not significantly different from titres of groups 79B or 71B’ for either PvDBPII SalI or PvW1. The only statistically significant differences were found for antibody titres for recognition of both PvDBPII SalI and PvW1 between groups 79A and 71B (Figure 1B). Control sera from volunteers that did not receive any immunization were seronegative for recognition of PvDBPII SalI and PvW1 (arbitrary units, AU ≤1) at all time points including C-2.

We also assessed the ability of anti-PvDBPII sera to block DARC-receptor binding by PvDBPII SalI and PvW1. Sera from control unvaccinated groups at C-2 and day 0 sera from volunteers collected prior to immunization showed DARC-binding inhibition of ≤30% for PvDBPII SalI and PvW1 at the lowest serum dilution of 1:10 that was tested in the binding inhibition assay. Binding inhibition for all vaccine groups at C-2 tested at 1:10 dilution was greater >90%. Anti-sera of 79A group with delayed boost of PvDBPII/Matrix-M^TM^ tended to have the highest binding inhibitory titres at C-2 (Figure 1B). However, only comparison between 79A and 71B reached statistical significance. There was no difference in binding inhibitory titres for PvDBPII SalI and PvW1 in all vaccinated groups at C-2.

In order to examine the stability of anti-PvDBPII antibodies, we analyzed correlations in antibody recognition titres and binding inhibitory titres at time points C-2, C+56 and C+96 (Supplementary Figure S2). The antibody titres and binding inhibitory titers were highly correlated between time points C-2 and C+56 or C+96.

### PvDBPII-specific antibodies can recognize native antigen in *P. vivax* schizonts

The ability of anti-PvDBPII sera to recognize native PvDBP in *P. vivax* schizonts was determined by IFA. All sera from vaccinated volunteers collected on day C-2 showed apical staining in merozoites within mature *P. vivax* schizonts (Figure 2A). Day 0 control sera collected prior to immunization (D0) did not show any specific signal in IFA with *P. vivax* schizonts (Figure 2A). Sera collected at C-2 from the 79A group had the highest reactivity at dilutions of 1:500 and 1:5000 (Figure 2B). Day C-2 sera from the 71B’ group had higher signal than sera collected at C-2 from groups 79B and 71B at dilution of 1:5000. Sera at C-2 from 79B group had stronger reactivity than 71B at 1:500 (Figure 2B). Based on signal intensity, we can order the groups from highest to lowest serum reactivity in IFA as follows: 79A > 71B’ > 79B > 71B.

**Figure 2.**
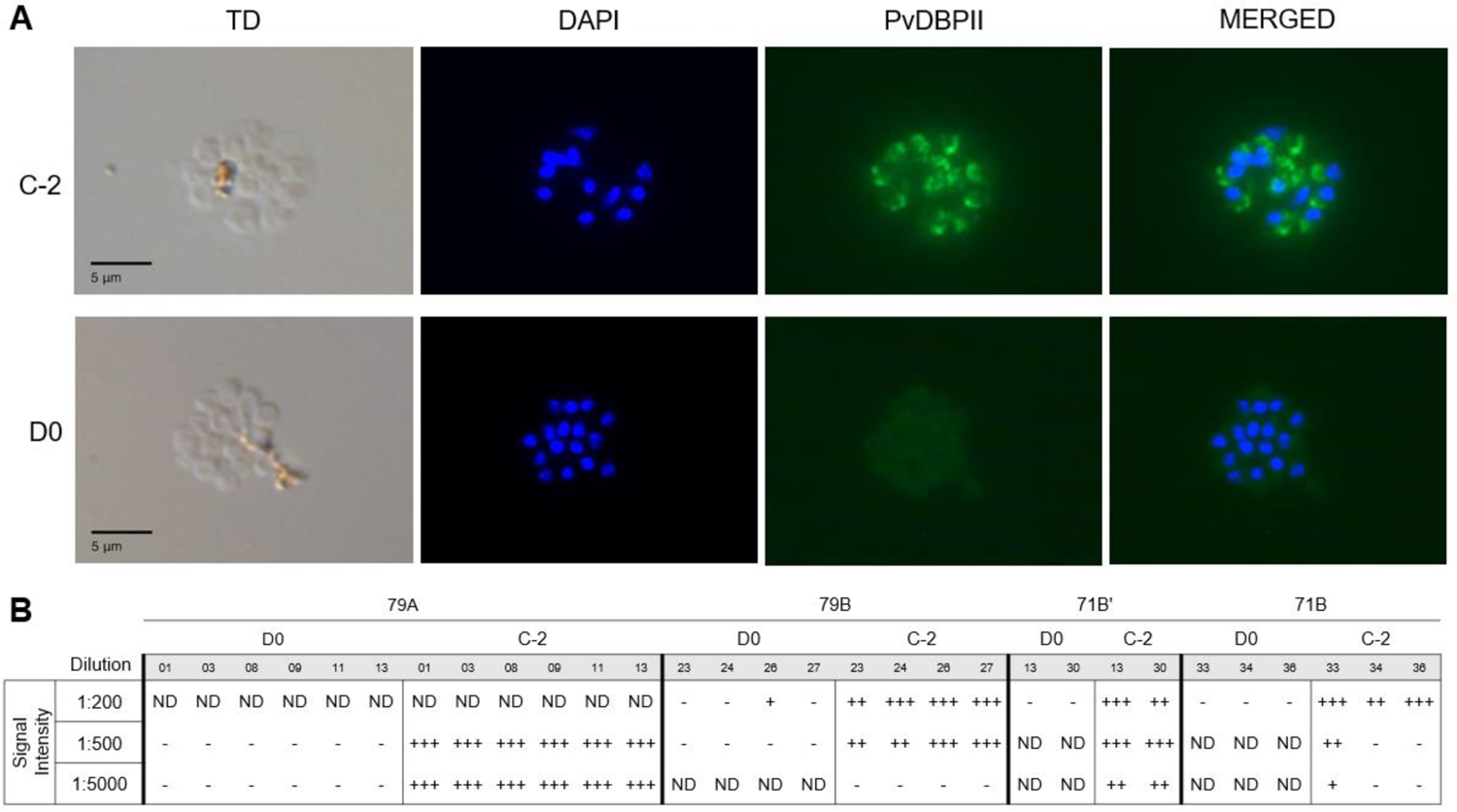
Reactivity of sera from immunized volunteers to *P. vivax* schizonts from infected *P. vivax* malaria patients. (**A**) Representative images of *P. vivax* schizonts incubated with sera from an individual of the 79A group at dilution 1:5000. Sera at time point C-2 showed apical staining to PvDBP (green) in *P. vivax* schizonts compared to sera collected prior to initial immunization (D0). (**B**) Table indicating the signal intensity in IFA of each of the individuals studied in the trials at C-2 and D0 and at different serum dilutions. The fluorescence intensity of each sample was scored as: +++, for high intensity; ++, medium intensity; +, low intensity; and -, no signal. Some IFA samples that were not done are designated as ND.

### Immune correlates for PMR reduction

To explore the immune processes that are associated with the PMR outcome, we measured diverse antibody functions and determined their correlation to PMR reduction calculated for each individual as: 100 – (test PMR x 100)/(PMR of unvaccinated controls). Multivariate analysis was performed with all the variables for both PvDBPII alleles SalI and PvW1 using the classification algorithm Boruta (24) to select the features that significantly contribute to the PMR outcome.

Sera from timepoint C-2 was used to evaluate i) the PvDBPII-binding kinetics of the polyclonal antibodies (affinity, association and dissociation constants or K_D_, k_a_ and k_dis_ respectively); ii) antibody isotypes and subclasses; iii) binding capacity for complement component C1q and Fc gamma receptors or FcγRs; iv) avidity; and v) ELISA recognition titres and binding inhibitory titres for PvDBPII. Both datasets corresponding to either antibody functions specific to PvDBPII SalI (Figure 3A) or to PvDBPII PvW1 (Figure 3C) showed significant correlations between most of the variables. We observed positive correlations between C1q binding, FcγR binding, isotypes, antibody recognition titres and binding inhibitory titres in both datasets (Figure 3A and 3C). Negative correlations were observed for the affinity and dissociation constants, K_D_ and k_dis_, in the PvDBPII SalI dataset (Figure 3A), whereas all three PvDBPII-antibody binding kinetic constants in the PvDBPII PvW1 dataset showed negative correlations with the rest of the dataset (Figure 3C). The IgG3 and IgM readouts did not show correlations to most of the features of both datasets (Figure 3A and 3C).

**Figure 3.**
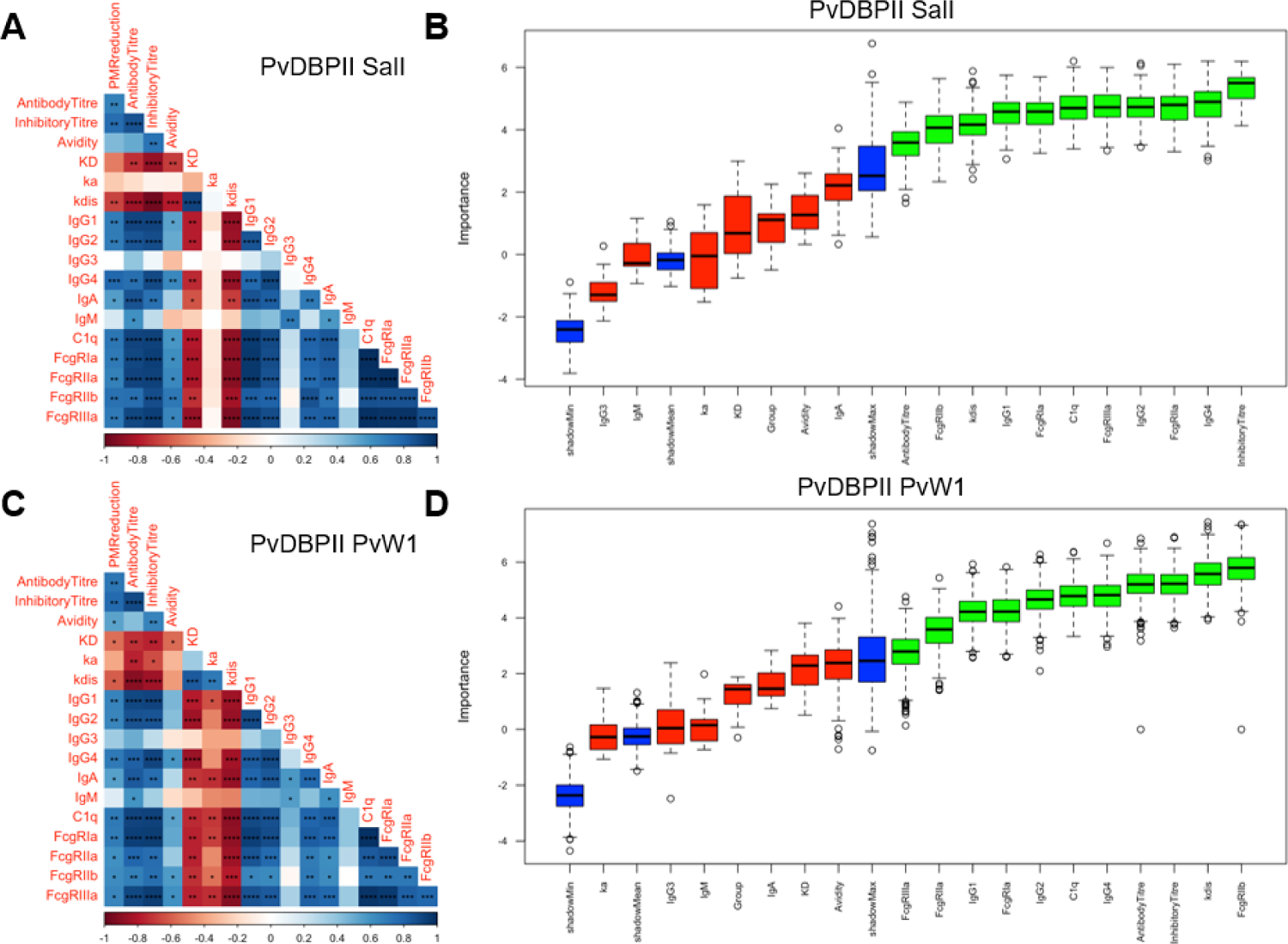
Variable correlations and feature selection of the antibody functions measured in the study. (**A**) Correlations between the anti-PvDBPII SalI antibody functions are shown. Correlation coefficients (shown in a double red-blue gradient) to each comparison were calculated using Spearman’s rank correlation tests. p values for each significant correlation are indicated, *p < 0.05 **p < 0.01 ***p < 0.001 ****p < 0.0001. (**B**) Importance plot for classification of variables specific to PvDBPII SalI that significantly contribute to the PMR reduction using the Boruta algorithm. The importance of each variable is defined as the Z-score of the mean decrease accuracy (normalised permutation importance). Blue boxes correspond to the minimal, average, and maximum Z-scores of shadow features. The variables that contribute significantly (green) or not (red) to the PMR reduction are shown. Boxplots show median Z-score (horizontal bar), interquartile range (boxes), range (whiskers), and outliers (open circles). (**C**) Correlations between the PvDBPII PvW1 antibody functions are depicted as in (**A**). (**D**) Importance plot for classification of variables specific to PvDBPII PvW1.

PMR reduction showed a significant correlation in the PvDBPII SalI dataset with antibody recognition titre, binding inhibitory titre, k_dis_, IgG1, IgG2, IgG4, IgA, C1q binding and FcγR binding (Figure 3A). In case of the PvDBPII PvW1 dataset, the PMR reduction showed significant correlations with the variables mentioned above for PvDBPII SalI dataset, K_D_ and avidity (Figure 3C). We proceeded to identify the variables that have a significant contribution to the PMR reduction outcome in both datasets. The Boruta algorithm selected as important the same 11 readouts in both datasets (Figure 3B and D). These important variables include binding inhibitory titres, FcγR binding and C1q binding, IgG1, IgG2, IgG4, k_dis_ and antibody titres. Binding inhibitory titre appeared as the variable with the highest importance in the PvDBPII SalI dataset (Figure 3B). In case of the PvDBPII PvW1 dataset, this variable also had high importance in the dataset (Figure 3D). Correlations of the PvDBPII SalI and PvW1 binding inhibitory titres to PMR reduction showed high significance (p ≤ 0.0014) (Figure 4A). This feature was also highly correlated when both PvDBPII alleles were compared (Figure 4B), indicating that immunization with PvDBPII SalI elicits antibodies that cross-react with PvDBPII PvW1. We modeled PMR reduction using a Random Forest regression with binding inhibitory titres specific to the two PvDBPII alleles. The importance of the two features as a PMR reduction predictor was similar and no statistical difference was observed (Figure 4C).

**Figure 4.**
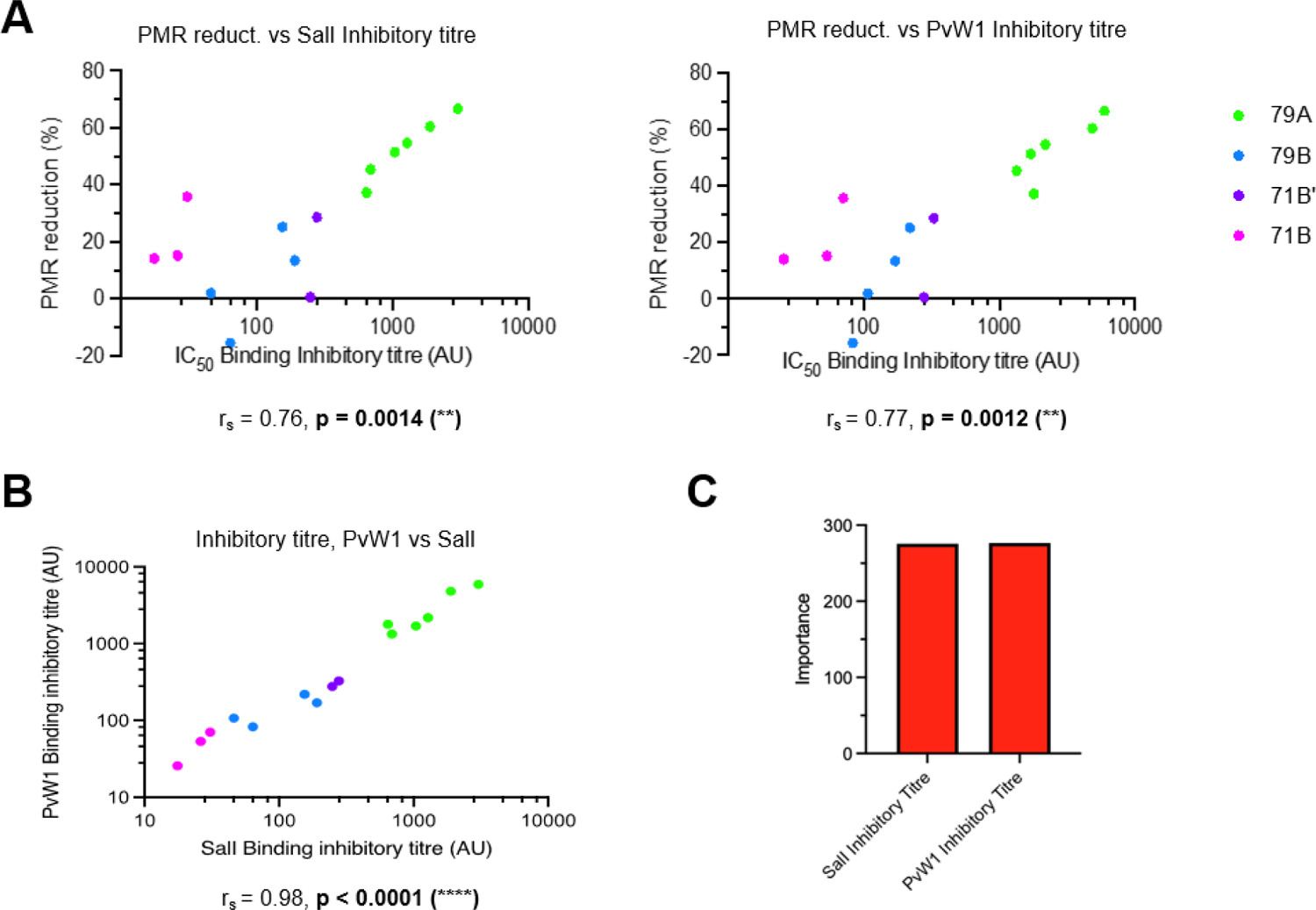
Binding inhibitory titre correlation to PMR reduction. (**A**) Individual correlations of the PMR reduction to binding inhibitory titres specific to PvDBPII SalI or PvW1. (**B**) Correlation between binding inhibitory titres specific to PvDBPII SalI and PvW1. (**A**) and (**B**) Correlations were calculated using Spearman’s rank correlation tests. Correlation coefficients and p values for each comparison are shown. (**C**) Importance plot of the binding inhibitory titres specific to PvDBPII SalI and PvW1 using Random Forest regression of the PMR reduction.

Correlations to PMR reduction for the rest of the variables considered as important by the Boruta algorithm in the PvDBPII SalI and PvW1 datasets are shown in Supplementary Figures S3 and S4, respectively. All these correlations to PMR reduction were statistically significant (Supplementary Figures S3 and S4). The unimportant variables of both datasets are also shown in Suplementary Figures S5 and S6 (PvDBPII SalI and PvW1 datasets, respectively). In case of the variables specific to PvDBPII SalI, IgA was significantly correlated to PMR reduction but it was not considered as an important predictor by the Boruta algorithm (Supplementary Figure S5). In case the variables specific to PvDBPII PvW1, avidity, K_D_ and IgA also correlated to the PMR reduction but they were not significant predictors as per the Boruta algorithm (Supplementary Figure S6). Next, we compared individually the 10 variables selected as important by the Boruta algorithm between the PvDBPII SalI and PvW1 datasets, with the exception of the binding inhibitory titres, already shown in Figure 4B. We observed significant correlations for all the variables (p ≤ 0.001) (Supplementary Figure S7), indicating strong cross-reactivity of polyfunctional PvDBPII-specific antibodies that correlate with PMR reduction.

## Discussion

Two vaccine platforms, recombinant protein-in-adjuvant and viral vectors were used to deliver the leading vivax malaria vaccine candidate PvDBPII to volunteers in a challenge trial to evaluate their efficacy (22). A PMR reduction of 53% was observed in the volunteers who received PvDBPII/Matrix-M^TM^ (79A group) in a delayed dosing regimen of 0, 1 and 14 months (22). No impact on the PMR was found in groups that received PvDBPII/Matrix-M^TM^ (79B group) with monthly regimen of 0, 1 and 2 months or viral-vectored PvDBPII at 0 and 2 months (71B group) or 0, 17 and 19 months (71B’ group) (22). Here, we performed exploratory analysis to evaluate the antibody responses in vaccinated volunteers to identify immune correlates associated with PMR reduction.

All vaccinated volunteers seroconverted after immunizations with both vaccine platforms. Sera collected 2 days before challenge (day C-2) was analyzed for polyfunctional antibody responses. Day C-2 sera from the 79A group tended to have higher anti-PvDBPII recognition titres, binding inhibitory titres and reactivity to native PvDBP in *P. vivax* schizonts by IFA compared to other groups. These findings are in accordance with studies showing improved antibody responses and neutralizing activity with delayed dosing regimens in vaccines targeting *P. falciparum* blood-stage antigen PfRH5 (25,26) and SARS-Cov-2 (27,28). This improved immunogenicity in delayed dosing regimens may arise from increased B cell populations. Indeed, the exploratory analyses of the PvDBPII-specific cellular responses conducted in the volunteers of this trial showed that memory B cells (CD19+ CD27+ IgG+) and plasma cells (CD19+ CD27+ CD38+) were significantly higher in delayed dosing regimens 7 days after final boost with both vaccine platforms used here (29). In addition, these two B cell populations are significantly correlated to PMR reduction (29). Extending the time between immunizations thus substantially improves antibody responses to PvDBPII potentially translating into greater efficacy.

The administration of the delayed dosing regimen of PvDBPII/Matrix-M^TM^ (79A group) resulted in a PMR reduction of 53% compared to unvaccinated controls. To our knowledge, this PMR reduction is the highest observed for any blood-stage malaria vaccine tested in CHMI (25,30–34). The highest PMR reduction observed in CHMI following immunization with a blood stage antigen thus far was 17% for the *P. falciparum* blood stage antigen, PfRH5 (25). Immunization with other *P. falciparum* blood stage antigens such as PfMSP1_42_ and PfAMA1 did not elicit significant parasite growth inhibition *in vivo* (30–34). A malaria vaccine should protect against infection by diverse strains. Immunization with the PvDBPII SalI variant elicits cross-reactive antibodies capable of inhibiting DARC-binding by multiple PvDBPII variants in pre-clinical (18,19) and clinical (20,21) studies. PvDBPII-specific antibodies inhibit binding to DARC by both PvDBPII alleles, SalI and PvW1, with similar efficiency (Figure 1B). The essential residues of PvDBPII that bind to DARC are conserved in the highly divergent PvW1 sequence (13–15). Additional analyses are required to determine if these binding inhibitory antibodies target these conserved DARC binding residues.

The immune correlates of protection against *P. vivax* are not clearly defined. Here, in addition to determining antibody recognition and binding inhibitory titres, we evaluated polyfunctional antibody responses elicited by PvDBPII SalI immunization to identify immune parameters that are associated with PMR reduction. We found that in addition to antibody recognition and binding inhibitory titres, PMR reduction is correlated with k_dis_, IgG1, IgG2, IgG4, IgA, C1q binding and FcγR binding for antibodies against both PvDBPII SalI and PvW1 variants (Figure 3A and 3C). Using the Boruta classifying algorithm, we obtained the same 11 variables that significantly contribute to PMR reduction in both PvDBPII SalI and PvW1 datasets (Figure 3B and 3D). Although a statistical analysis cannot definitively prove that a biomarker is causally associated with protection rather than just correlated, the variable that showed the highest importance in the PvDBPII SalI dataset was the binding inhibitory titre (Figure 3B), which also had high importance in the PvDBPII PvW1 dataset (Figure 3D). The binding inhibitory titres specific to the two PvDBPII variants were highly correlated (Figure 4B) and modeled PMR reduction similarly when combined (Figure 4C). This may indicate the significant value of PvDBPII-DARC binding inhibitory titres as a *P. vivax* protection predictor. The high correlation between PvDBPII binding inhibitory titres for SalI and PvW1 (Figure 4B) reflects the cross-reactivity of these high titre binding inhibitory antibodies. Children residing in a malaria-endemic area of Papua New Guinea (PNG), who developpedc high-titre cross-reactive binding inhibitory antibodies to PvDBPII had a reduced risk of *P. vivax* infection and lower parasite densities (16). However, the IC_50_ binding inhibition titres of these naturally acquired antibodies was ∼1:20 (16). In comparison, day C-2 sera of 79A group showed 50-100 times higher binding inhibition titres with IC_50_ of ∼1:1000 to 1:2000 (Figure 1B). Given that PvDBPII immunization elicits substantially higher binding inhibitory titres compared to naturally acquired binding inhibitory antibodies that are associated with protection, one can expect immunization-induced antibodies to protect against *P. vivax* infection. The *P. vivax* blood-stage challenge model allows one to determine if a vaccine elicits antibodies that have a biological effect to reduce parasite growth *in vivo*. Whether the PMR reduction of 53% (22) translates to protection against clinical disease remains to be determined. Protection against clinical *P. vivax* malaria will need to be determined in field trials in a malaria-endemic area.

In addition to binding inhibitory antibodies, ELISA recognition titres and presence of anti-PvDBPII-specific IgG1, IgG2 and IgG4 significantly contributed to PMR reduction. Consistent with our results, total IgG as well as IgG1 specific to PvDBPII SalI were associated with a reduced risk of malaria in exposed children from PNG (35). In addition, the He et al. study (35) reported undetectable IgG3 levels for PvDBPII SalI similar to the very low levels of IgG3 detected here (Supplementary Figure S5). However, IgG3 against PvDBPII allele AH, the most frequent allele in this region, was detected in the PNG field study (35). IgG3 against PvDBPII allele AH was more predominant in older children and correlated with protection (35). Other studies in malaria-endemic areas showed that there is a higher prevalence of IgG3 specific to merozoite surface proteins in *P. vivax* (36) and *P. falciparum* (37,38) with age. This evidence suggests that class-switching to antigen-specific IgG3 may develop after multiple *P. vivax* infections but not after a limited number of immunizations with PvDBPII. The IgG2 and IgG4 subclasses, which are rarely found upon natural exposure to both *P. falciparum* and *P. vivax* infections, were significant predictors of PMR reduction according to our feature selection analysis. These IgG subclasses do not bind FcγRs as efficiently as IgG1 and IgG3 (39). The immune effector mechanisms induced by anti-PvDBPII specific IgG2 and IgG4 antibodies remain to be identified.

The binding of anti-PvDBPII antibodies to FcγRs was also a significant predictor of PMR outcome. The cellular responses mediated by these receptors could play an important role in parasite growth reduction and protection against clinical symptoms as seen for *P. falciparum*. After a Phase II trial showing that vaccination with RTS,S (based on the *P. falciparum* circumsporozoite protein, PfCSP) protects against *P. falciparum* infection in malaria-naïve volunteers (40), multivariate analysis revealed that FcγRIIIa and antibody-dependent cellular phagocytosis (ADCP) were the strongest correlates of protection in a sporozoite-based CHMI (41). Binding of anti-PfCSP antibodies to C1q was also found to correlate with RTS,S-elicited protection (41). Largely unknown in *P. vivax*, complement-fixing antibodies of diverse blood-stage antigens in *P. falciparum* have been correlated with protection upon natural exposure (42). Our data suggests that complement and FcγRs may also contribute to *P. vivax* growth reduction.

Importantly, Suscovich and collaborators found that IgA is a highly important predictor of RTS,S-induced protection (41). Similarly, multivariate analysis of immune responses in a Phase I/IIa trial with blood-stage *P. falciparum* vaccine PfRH5 revealed that IgA responses were the variables with the greatest importance that contribute to PMR reduction in malaria-naïve individuals (25). Contrary to this, our data suggest that anti-PvDBPII IgA was not a significant predictor even though it was individually correlated to PMR reduction (Supplementary Figures S5 and S6). In addition, avidity and isotype IgM did not significantly contribute to PMR reduction. After volunteers were immunized with RTS,S and underwent CHMI, IgM was found to be a susceptibility marker for parasite infection as it was more predominant in infected volunteers (41). In case of avidity, it also increased in malaria-naïve volunteers who received the delayed third dose of PfRH5 compared to a monthly dosing regimen (25). We observed a similar trend of higher avidity of anti-PvDBPII antibodies in delayed dosing regimens compared to shorter dosing schedules of both vaccine platforms (79A vs 79B and 71B’ vs 71B). However, our feature selection algorithm showed that avidity was not an important variable for *P. vivax* PMR reduction unlike the case for anti-PfRH5 avidity and *P. falciparum* (25). In addition, two field trials analyzing the vaccine efficacy of RTS,S in children found that anti-PfCSP avidity was not statistically associated with protection (43,44).

In terms of binding kinetics of PvDBPII and anti-PvDBPII antibodies, the affinity constant, K_D_, and association constant, k_a_, were not selected as important by the feature selection algorithm in both datasets. Instead, the algorithm identified that the dissociation constant, k_dis_, was a significant contributor to PMR reduction. The k_dis_ for anti-PvDBPII polyclonal antibodies indicates the stability of antibodies bound to PvDBPII. The ability of anti-PvDBPII antibodies to remain bound to PvDBPII could lead to better neutralizing and effector activities resulting in significant PMR reduction.

The data presented here identify key functional properties of PvDBPII-specific antibodies elicited by immunization that predict PMR reduction following *P. vivax* blood-stage challenge. PvDBPII-binding inhibitory titres showed high importance for prediction of PMR reduction, though other features such as antibody titre, k_dis_, IgG1, IgG2, IgG4 and binding to C1q and FcγRs may also contribute to *P. vivax* protection.

PvDBPII/Matrix-M^TM^ delivered in a delayed dosing regimen has shown significant reduction in PMR. Extensive analysis of immune responses reported here has identified for the first time functional antibody immune correlates. These immune correlates together with anti-PvDBPII binding inhibitory titres should be evaluated in subsequent trials and could guide the clinical development of a high efficacy *P. vivax* vaccine based on PvDBPII.

## Material and Methods

### Study design

A full description of the Phase I/IIa clinical trial is described elsewhere (22). Serum samples were obtained from a series of three Phase I/IIa clinical trial protocols (called VAC069, VAC071 and VAC079) conducted in parallel and designed to evaluate the efficacy of immunization with PvDBPII vaccine candidates including His-tag free recombinant PvDBPII SalI formulated with Matrix-M^TM^ adjuvant and viral-vectored PvDBPII SalI delivered by ChAd63 and MVA followed by blood stage challenge with the Thai *P. vivax* clinical isolate PvW1 to evaluate efficacy (22).

### Expression of recombinant PvDBPII and DARC-Fc

Recombinant PvDBPII SalI (the vaccine candidate) and PvDBPII PvW1 (the binding domain of PvDBP from the *P. vivax* isolate PvW1 used for CHMI) were produced as previously described (19,45–47). Briefly, synthetic genes encoding PvDBPII SalI and PvW1 with C-terminal 6-His tags that were codon optimized for expression in *E. coli,* were cloned into the pET28a (+) vector (GenScript) and the resultant plasmids were transformed into *E. coli* strain BL21(DE3) pLysS (C6060, Thermo Fisher). PvDBPII was expressed by fed-batch fermentation, cells were lysed, PvDBPII was solubilized from inclusion bodies under denaturing conditions and purified by nickel-charged nitrilotriacetic acid (Ni-NTA) affinity chromatography (17524802, Cytiva). Recombinant PvDBPII was refolded by rapid dilution method, dialyzed and finally purified by cation exchange (SP Sepharose column, 17115201, Cytiva) and gel filtration (Superdex 200 column) chromatography (28-9893-35, Cytiva). The final monomeric recombinant PvDBPII was stored at −80°C.

A plasmid encoding DARC-Fc was generated by ligating the first 60 codons of human DARC (FyB allele) to sequences encoding the Fc region of human IgG1 in the mammalian expression vector pCDM8 (48). The plasmid containing the human tyrosylprotein sulfotransferase-2 (TPST-2) (48) was co-transfected with the DARC-Fc plasmid into HEK293T cells (CRL-3216, ATCC). Recombinant DARC-Fc was purified from culture supernatants by protein G affinity chromatography. The final recombinant protein was stored at −80°C.

### ELISA

Nunc MaxiSorp ELISA plates (439454, Thermo Fisher) were coated overnight with 100μl of recombinant PvDBPII variant SalI or PvW1 proteins (1 μg/ml) in carbonate-bicarbonate buffer (C3041, Sigma) at 4°C. Next day, plates were washed three times with 0.05% Tween (P1379, Sigma) in PBS (PBS/T) and blocked with 200 μl of 5% non-fat milk (Regilait) PBS/T for 1 h at 37°C. Test and reference anti-PvDBPII sera were diluted in 2.5% milk (initial dilution 1:200) and 100 μl was added per well in duplicate and incubated for 1 h at 37°C. After washing, bound antibodies were detected by adding 100 μl of horseradish peroxidase-conjugated anti-human IgG rabbit antibodies (A8792, Sigma) at dilution 1:4000 and incubated for 1 h at 37°C. The assay was developed at room temperature by using 100 μl of the two-component chromogenic substrate for peroxidase detection, TMB (3,3′,5,5′-tetramethylbenzidine, 5120-0047, Life Sciences), for 5 min and the reaction was stopped with 100 μl of 1 M phosphoric acid (H_3_PO_4_, 695017, Sigma). The optical density was measured immediately at a wavelength of 450 nm (OD_450_). The reference serum was assigned 200 arbitrary units, AU, and the standard curve from the reference serum was used to fit a four-parameter logistic model using ADAMSEL version 3.0 software (Ed Remarque© 2021). OD_450_ values were later converted to AU using the standard curve and antibody titres of test sera were reported in AU for each sample.

### Avidity

PvDBPII SalI or PvW1 pre-coated ELISA plates were incubated with test samples diluted to give an OD_450_ of ∼2.0. After sample incubation, descending concentrations of the chaotropic agent sodium thiocyanate (NaSCN, 251410, Sigma) (7 M to 0 M in PBS) were added (100 μl) and incubated for 15 min at room temperature. Plates were washed with PBS/T and reaction was developed to detect bound IgGs as per ELISA protocol. OD_450_ values were plotted versus NaSCN concentration and fitted in a four-parameter logistic model. The NaSCN concentration that resulted in a 50% reduction of the OD_450_ was used as a measure of the avidity (IC_50_).

### Isotyping

PvDBPII SalI or PvW1 pre-coated ELISA plates were incubated with test samples diluted 1:100 in duplicates. After washing, the following antibodies were added for detection (100 μl at dilution 1:1000): biotin-conjugated mouse monoclonal anti-human IgG1 (B6775, Life Technologies); biotin-conjugated mouse monoclonal anti-human IgG2 (B3398, Life Technologies); biotin-conjugated mouse monoclonal anti-human IgG3 (B3623, Sigma); biotin-conjugated mouse anti-human IgG4 (B3648, Sigma); peroxidase-conjugated goat polyclonal anti-human IgA α-chain (A0295, Sigma); and peroxidase-conjugated goat polyclonal anti-human IgM μ-chain (401905, Millipore). After 1 h incubation at 37°C, plates were washed and avidin-peroxidase (A7419, Sigma) was added (100 μl at dilution 1:5000), except for the IgA and IgM wells, to which 2.5% milk was added. After 1 h incubation at 37°C, the reaction was developed like in ELISA protocol. The OD_450_ was used to evaluate the IgG subclass, IgA or IgM of each sample.

### ELISA-based PvDBPII-DARC Binding Inhibition Assay

The binding of PvDBPII to DARC was analyzed in presence or absence of anti-PvDBPII antibodies using an ELISA based format as described earlier (48). Briefly, recombinant DARC-Fc (1 μg/ml) was coated on to Nunc MaxiSorp ELISA plates overnight at 4°C in carbonate-bicarbonate buffer. Next day, the plate was blocked for 2 h at 37°C using 2% non-fat milk PBS/T. Recombinant PvDBPII SalI or PvW1 with concentrations in the range of 0.8-25 ng/ml were used to generate the PvDBPII standard curve using a four-parameter logistic curve. Serum samples were analyzed at dilutions 1:10 to 1:2430. Each serum dilution was incubated with 25 ng/ml PvDBPII at 37°C for 30 min. The reaction mixture was added to wells in duplicate and incubated at 37°C for 1 h. PvDBPII bound to DARC was probed with anti-PvDBPII polyclonal rabbit sera (generated in house) at 37°C for 1 h and detected with peroxidase-conjugated anti-rabbit IgG secondary antibody (A6154, Sigma) at 37°C for 1 h. The assay was developed as described in ELISA protocol. The amount of bound PvDBPII was estimated by converting OD_450_ values to protein concentrations using the PvDPBII standard curve. The interpolated protein concentration values were used to calculate percent binding (%) for each serum sample dilution. Then, the % binding inhibition for each serum dilution was calculated as follows: % Binding Inhibition = 100 - % Binding. The plot of % Binding Inhibition versus serum dilution was used to find the serum dilution at which 50% binding inhibition (IC_50_) is achieved. Three independent replicates were averaged to determine the median IC_50_.

### Immunofluorescence assay (IFA)

Frozen slides of *P. vivax* schizonts were thawed at room temperature for 30 min. Slides were blocked with 5% bovine serum albumin (BSA, A7030, Sigma) in PBS for 30 min at 37°C and probed with test sera diluted in 2.5% BSA at 1:200, 1:500 and 1:5000 for 30 min at 37°C, followed by three washes with PBS. A mixture of Alexa Flour 488-conjugated goat anti-human IgG (H+L) secondary antibodies (A11013, Themo Fisher) at 1:500 and Hoeschst 33342 solution (62249, Thermo Fisher) at 1:20000 was added and incubated for 30 min at 37°C. After washing, slides were treated with anti-Fade (H-1000-10, Vector Laboratories) and visualized on Leica DM 5000B Microscope (Leica Microsystems). The fluorescence intensity of each sample was scored as follows: +++, for high intensity; ++, medium intensity; +, low intensity; and -, no signal.

### Antibody affinity by biolayer interferometry (BLI)

Experiments were performed on an Octet RED 384 instrument (Fortebio) at 25°C with shaking at 1000 round per minute (rpm). All assays were conducted in standard Greiner black 96-well microtiter plates (655209, Greiner) in a volume of 120 μl/well. Buffer consisting of PBS with 1 mg/ml BSA was used for baselines, dissociation steps and to dilute recombinant proteins. Sera were diluted in PBS to achieve final concentrations of IgGs in the range of 30 to 0 nM. The method was set as follows: NTA biosensors (18-5101, Sartorius) were hydrated for 10 min in PBS and regenerated (3 cycles, 30 s each) with 10 mM glycine pH 1.5 (410225, Sigma), followed by another regeneration with 50 mM ethylenediaminetetraacetic acid (EDTA) (EDS, Sigma). Biosensors were activated with 10 mM nickel sulfate (NiSO_4_, 656895, Sigma) for 180 s and later dipped in buffer (120 s). PvDBPII variant SalI or PvDBPII PvW1 were immobilized via His tag at 5 μg/ml for 600 s. A mammalian cytosolic protein, peroxiredoxin 6 (PRDX6) (49), was loaded at 3 μg/ml to pre-charged NTA biosensors for 600 s as a reference biosensor (negative control). Loaded biosensors were tested for binding to test sera in the following steps: baseline (60 s in buffer), association step (600 s in serum dilutions) and dissociation step (600 s in buffer). Two wells containing only buffer instead of sera were assigned as reference wells. Signals from reference wells and reference biosensors were later subtracted. Affinity, association and dissociation constants (K_D_, k_a_ and k_dis_, respectively) were calculated by Data Analysis HT software version 10 (Fortebio) using kinetic analysis and a 1:1 binding model.

### Binding analysis to complement component C1q and Fc receptors by BLI

Experiments were performed on an Octet HTX instrument (Fortebio) at 25°C with shaking at 1000 rpm. All assays were conducted in standard black 384-well microtiter plates (781209, Greiner) and volume was 120 μl/well. The following recombinant proteins diluted in PBS containing 1 mg/ml BSA (PBS-BSA) were used to determine binding to sera: C1q (ab282858, Abcam) at 12 nM; the Fc gamma receptor (FcγR) Ia (FcγRIa), FcγRIIa, FcγRIIb, and FcγRIIIa (all from Sino Biological: 10256-H08H, 10374-H08H1, 10259-H08H and 10389-H08H1, respectively) at dilutions 19 nM, 1.7 μM, 3.7 μM and 2.3 μM, respectively. The method was set as follows, including the reagents from the AR2G Reagent kit (18-5095, Sartorius): Amine-reactive second-generation (AR2G) biosensors (18-5092, Sartorius) were activated as per manufacturer’s protocol in 20 mM (1-Ethyl-3-(3-Dimethylamino propyl) Carbodiimide, hydrochloride (EDC) and 10 mM sulfo N-hydroxysuccinimide (s-NHS) solution for 300 s. PvDBPII SalI domain, PvDBPII PvW1 recombinant protein or PRDX6 (reference biosensor) were immobilized at 20 μg/ml in 10 mM acetate buffer pH 6 for 600 s. Loaded biosensors were quenched with 1 M ethanolamine pH 8.5 for 300 s. Later, biosensors were regenerated (3 cycles, 30 s each) with 10 mM glycine pH 1.5 and dipped into wells containing serum diluted in PBS-BSA at 1:100 for 600 s. After an incubation of 900 s in PBS-BSA to stabilize the signals, biosensors were tested for binding to C1q and FcγRs in the following steps: baseline (120 s in PBS-BSA), association step (600 s in C1q and FcγR dilutions) and dissociation step (600 s in PBS-BSA). Wells containing PBS-BSA alone were assigned as reference wells. Signals from reference wells and reference biosensors were later subtracted. Binding to C1q and FcγRs was measured as the wavelength shift signal or response (nm) at the end of the association step. All the data were analyzed using Octet BLI Analysis Studio 13.0 software.

### Statistics

Comparisons among vaccinated groups and timepoints in antibody and binding inhibitory titres were performed using a Kruskal–Wallis test with Dunn’s correction for multiple comparisons. Differences between SalI and PvW1 were tested with pairwise comparisons with Bonferroni’s multiple comparison tests. Correlations between variables were determined by Spearman rank test. Modeling of PMR reduction including only the binding inhibitory titres specific to both PvDBPII alleles was performed by Random Forest regression. All statistical tests were two-sided and a p-value < 0.05 was considered significant. Data analysis and graphs were performed using GraphPad Prism version 9.3.1 (GraphPad Software Inc.). The correlation plot of all variables and Random Forest regression were generated in R version 4.2.2 and RStudio version 2022.12.0+353.

Feature selection was performed using a Boruta algorithm (24,50). The Boruta algorithm is a wrapper method built around a random forest classifier that performs a top-down search for relevant features, while progressively eliminating irrelevant features, by comparing the importance of original features with the importance achievable at random (shadow features, estimated using permuted copies of the original features). To measure the importance of the immunological readouts that significantly contribute to the PMR outcome, we applied the Boruta algorithm using R version 4.2.2 and RStudio version 2022.12.0+353.

## Ethics Statement

Sera from volunteers participating in the three clinical trials conducted at the University of Oxford, UK were sent to Institut Pasteur, Paris under a Material Transfer Agreement. The clinical trial studies had received ethical approval from UK National Health Service Research Ethics Services, (VAC069: Hampshire A Research Ethics Committee, Ref 18/SC/0577; VAC071: Oxford A Research Ethics Committee, Ref 19/SC/0193; VAC079: Oxford A Research Ethics Committee, Ref 19/SC/0330). The vaccine trials were approved by the UK Medicines and Healthcare products Regulatory Agency (VAC071: EudraCT 2019-000643-27; VAC079: EudraCT 2019-002872-14). The trials are registered under the following ClinicalTrials.gov numbers: VAC069 NCT03797989, VAC071 NCT04009096, VAC079 NCT04201431. The research conducted at the Institut Pasteur received approval from the Institutional Review Board (IRB) of the Institut Pasteur (Research Ref IRB2022-03).

## Data Availability

All data that support the findings of this study will be made available online with the manuscript.

## Acknowledgements

Development of PvDBPII as a vaccine candidate was supported by grants from the Biotechnology Industry Research Assistance Council (BIRAC), New Delhi and PATH Malaria Vaccine Initiative. MVDP was supported by grants from the Bill and Melinda Gates Foundation and Department of Biotechnology (DBT), Government of India. This work was also supported in part by grants from Agence Nationale de Recherche to CEC [ANR-18-CE15-0026 and ANR-21-CE15-0013-01]. CEC is supported by the French Government’s Laboratoire d’Excellence “PARAFRAP” [ANR-11-LABX-0024-PARAFRAP] in addition to core funding from Institut Pasteur. FJM was supported by a Fellowship from Ecole Doctorale BioSPC (ED562), F-75006, Université Paris Cité.

The authors are grateful for the assistance of the VAC069, VAC071 and VAC079 clinical trial teams and all the study volunteers. The VAC069 and VAC071 trials were funded by the European Union’s Horizon 2020 research and innovation program under grant agreement 733073 for MultiViVax. The VAC079 trial was funded by the Wellcome Trust Malaria Infection Study in Thailand (MIST) program [212336/Z/18/Z]. For the purpose of open access, the author has applied a CC BY public copyright licence to any Author Accepted Manuscript version arising from this submission. This work was also supported in part by the UK Medical Research Council (MRC) [G1100086] and the National Institute for Health Research (NIHR) Oxford Biomedical Research Centre (BRC). The views expressed are those of the authors and not necessarily those of the NHS, the NIHR or the Department of Health. CMN held a Wellcome Trust Sir Henry Wellcome Postdoctoral Fellowship [209200/Z/17/Z]. SJD is a Jenner Investigator and held a Wellcome Trust Senior Fellowship [106917/Z/15/Z].

## Author contributions

FJM expressed recombinant proteins, performed assays, analyzed data and wrote the manuscript. MW analyzed data. MGB performed immuno-assays. CH expressed recombinant proteins. AB, FA and PE reviewed data. JP provided samples. MMH, SES, JRB, CMN generated and provided samples. JMR provided Matrix-M^TM^ adjuvant. PM and VSC supported vaccine development, production and project management. AMM and SJD were investigators on the clinical trial. CEC developed the PvDBPII/Matrix M vaccine, designed the study, analyzed the data and wrote the manuscript. All authors reviewed the manuscript and agreed with the final version.

## Declarations of Interest

SJD has provided consultation services to GSK on malaria vaccines, is an inventor on patent applications relating to adenovirus-based vaccines and is an inventor on intellectual property licensed by Oxford University Innovation to AstraZeneca. AMM has provided consultation services to GSK on malaria vaccines and has an immediate family member who is an inventor on patents relating to adenovirus-based vaccines, and is an inventor on intellectual property licensed by Oxford University Innovation to AstraZeneca. CEC is an inventor on patents that relate to binding domains of erythrocyte-binding proteins of Plasmodium parasites including PvDBP. JMR is an employee of Novavax, developer of the Matrix-M^TM^ adjuvant.

## Notes

### Clinical Trial

The trials are registered under the following ClinicalTrials.gov numbers: VAC069 NCT03797989, VAC071 NCT04009096, VAC079 NCT04201431. The research conducted at the Institut Pasteur received approval from the Institutional Review Board (IRB) of the Institut Pasteur (Research Ref IRB2022-03).

### Author Declarations

The clinical trial studies had received ethical approval from UK National Health Service Research Ethics Services, (VAC069: Hampshire A Research Ethics Committee, Ref 18/SC/0577; VAC071: Oxford A Research Ethics Committee, Ref 19/SC/0193; VAC079: Oxford A Research Ethics Committee, Ref 19/SC/0330). The vaccine trials were approved by the UK Medicines and Healthcare products Regulatory Agency (VAC071: EudraCT 2019-000643-27; VAC079: EudraCT 2019-002872-14). The trials are registered under the following ClinicalTrials.gov numbers: VAC069 NCT03797989, VAC071 NCT04009096, VAC079 NCT04201431. The research conducted at the Institut Pasteur received approval from the Institutional Review Board (IRB) of the Institut Pasteur (Research Ref IRB2022-03).

